# Genome-wide Association Studies of Missing Metabolite Measures: Results From Two Population-based Studies

**DOI:** 10.1101/2024.10.02.24314800

**Authors:** Tariq O. Faquih, Mohammed Aslam Imtiaz, Valentina Talevi, Elvire N. Landstra, Astrid van Hylckama Vlieg, Ruifang Li-Gao, Frits R. Rosendaal, Raymond Noordam, Diana van Heemst, Dennis O. Mook-Kanamori, Monique M. B. Breteler, N. Ahmad Aziz, Ko Willems van Dijk

**Affiliations:** Department of Clinical Epidemiology, Leiden University Medical Center, Leiden, The Netherlands; Division of Sleep and Circadian Disorders, Brigham and Women’s Hospital, Boston, United States; Population Health Sciences, German Centre for Neurodegenerative Diseases (DZNE), Bonn, Germany; Department of Internal Medicine, Section of Gerontology and Geriatrics, Leiden University Medical Center, Leiden, The Netherlands; Department of Public Health and Primary Care, Leiden University Medical Center, Leiden, The Netherlands; Institute for Medical Biometry, Informatics and Epidemiology (IMBIE), Faculty of Medicine, University of Bonn, Bonn, Germany; Department of Neurology, Faculty of Medicine, University of Bonn, Bonn, Germany; Department of Human Genetics, Leiden University Medical Center, Leiden, The Netherlands; Department of Internal Medicine, Division of Endocrinology, Leiden University Medical Center, Leiden, The Netherlands; Einthoven Laboratory for Experimental Vascular Medicine, Leiden University Medical Center, Leiden, The Netherlands

**Keywords:** genome-wide association study, missing measurements, inborn errors of metabolism, metabolomics, poor metabolizers

## Abstract

Metabolomic studies are increasingly used for both etiological and predictive research, but frequently report missing values. We hypothesized that interindividual genetic variation may account for part of this missingness. Therefore, we performed a GWAS of missingness in measured metabolite levels using an untargeted mass spectrometry-based platform in the Netherlands Epidemiology of Obesity Study (N=594) and the Rhineland Study (N=4,165). We considered metabolites missing in 10%-90% of individuals in both cohorts (N=224). GWAS meta-analyses of these metabolites’ probability of missingness revealed 55 metabolome-wide significant associations, including 42 novel ones (p<1.58×10^-10^), involving 28 metabolites and 41 lead SNPs. Despite considerable pleiotropy, the majority of identified SNP-‘missing metabolite’ associations were biologically plausible, relating to beta-oxidation, bile acids, steroids, and xenobiotics metabolism. These findings suggest that missing values in metabolomics are nonrandom and partly reflect genetic variation, accounting for which is important for both clinical and epidemiological studies, especially nutritional and pharmacogenetics studies.

## Introduction

Metabolites are small molecules that are produced or consumed during anabolic or catabolic reactions and constitute the basic building blocks of all biological processes. Circulating metabolite levels are thought to reflect the integrated metabolic response to changes in genetic and non-genetic (including dietary and other environmental) factors.^1^ This hypothesis has made metabolomics an attractive field of study for elucidating the biological mechanisms underlying complex multifactorial diseases.^1,2^ Recent advances in metabolomics have enabled high-throughput analysis of thousands of metabolites from a single biological sample, and have been applied to study a wide range of cardiovascular,^3,4^ metabolic,^5,6^ and neurodegenerative outcomes,^7,8^ as well as other traits.^9–11^

The field of metabolomics remains relatively new and still faces several challenges. One important challenge is the biological meaning of missing measurements of metabolites, particularly with untargeted approaches.^12,13^ Conceptually, missing data could be due to either random or systematic (i.e., technical) measurement errors, or reflect the actual absence of specific metabolites. In addition, when the metabolite concentration in the sample is below the limit of detection of the measurement method, it will be reported as a missing value.^12,13^ Indeed, in most studies, missing data are assumed to reflect values below the limit of detection, and consequently are either removed from the analysis or imputed.^12,13^ However, *a priori*, it cannot be excluded that missing values of metabolites are caused by genetic variants. In this case, the metabolites with missing values could be truly absent from the sample due to functional alterations of specific biological pathways driven by genetic variation.^9,14,15^ Therefore, imputation or removal of those metabolites from the analysis could bias biological interpretation.

Long before large-scale metabolomics data became available, rare genetic mutations affecting metabolism were identified and investigated.^16^ Disorders caused by genetic mutations that disrupt metabolism are referred to as inborn errors of metabolism (IEM). Usually, the causal genetic mutations are located in protein coding genes and affect the structure of the encoded proteins to such an extent that their biological function is disrupted.^17^ For example, IEM disorders can disrupt carbohydrate metabolism, protein metabolism, fatty acid oxidation, and glycogen storage.^17,18^ Collectively, IEM disorders have an overall incidence of 1 in 2500 births.^18^ IEM illustrate that certain genetic variants have the potential to prevent the synthesis or breakdown of specific metabolites by disrupting metabolic pathways.^19^ We set out to test the hypothesis that at least some of the common missing values in metabolomics data, either due to levels below the limit of detection or otherwise, is caused by common genetic variation. We also hypothesized that the nature and context of the potential associations could provide insights into the potential causes of the missingness (i.e., technical, below limit of detection, or truly absent). To address these hypotheses, we performed genome-wide association studies (GWAS) to discover SNPs associated with the probability of absence (i.e., ‘missingness’ due to concentrations below the limit of detection or truly absent) of metabolite measures.

## Methods

### Study Populations

We included 594 and 4,165 individuals of European ancestry from the Netherlands Epidemiology of Obesity (NEO) study and the Rhineland Study, respectively, who had both genetic and metabolomics data. NEO is a population-based, prospective cohort study, initiated in 2008. All participants recruited in this study gave written informed consent and the Medical Ethical Committee of the Leiden University Medical Center (LUMC) approved the study design. A detailed description of the study design and data collection can be found elsewhere.^20^ Briefly, men and women aged between 45 and 65 years with a self-reported body mass index (BMI) of 27 kg/m^2^ or higher living in the greater area of Leiden (in the west of the Netherlands) were eligible to participate in NEO. Participants were invited for a baseline visit at the NEO center in the LUMC after an overnight fast. At the baseline visit, fasting blood samples were drawn. The Rhineland Study is an ongoing prospective population-based cohort study in Bonn, Germany. People aged 30 years or above who lived in two geographically defined areas in Bonn were invited to participate with the only exclusion criterium being insufficient command of the German language to provide informed consent. These participants underwent deep phenotyping to obtain whole-blood, genetic, imaging, socio-demographic, and clinical data.

### Metabolomics Measurements and Missingness Inclusion Criteria

Metabolites were measured on the Metabolon HD4 platform in the fasting state serum samples (N= 594) from NEO and fasting state plasma samples (N= 4,165) from the Rhineland Study. Details on the metabolomics pipeline have been described elsewhere.^11^ In brief, the Metabolon HD4 platform employs an untargeted measurement approach that utilizes ultra-performance liquid chromatography (UPLC) tandem mass spectrometry (MS/MS) combined with a positive ion mode electrospray ionization, RP/UPLC/-MS/MS combined with a negative ion mode electrospray, and HILIC/UPLC-MS/MS combined with a negative ion mode electrospray ionization. In total, 1,365 metabolites were measured in NEO and 1,077 were measured in the Rhineland Study. Of these, 847 and 467 were endogenous in NEO and the Rhineland Study, respectively. Based on the pathway annotations by Metabolon, these endogenous metabolites spanned 10 pathway groups: amino acids, cofactors and vitamins, lipids, energy, nucleotides, peptides, carbohydrates, and partially characterized molecules. In addition, it included measurements of 222 and 321 xenobiotic metabolites as well as 296 and 289 unannotated metabolites from NEO and the Rhineland Study, respectively.

Most of missingness for endogenous metabolites measured by the metabolon platform occurs in less than 10% of the measured population. These cases are commonly due to systematic or random errors in measurements. On the other hand, for xenobiotic metabolites most missing values are found in 90% of the population as these metabolites depend on specific external exposures (i.e., medication use, nicotine exposure etc.). Therefore, we included metabolites with a moderate number of missing values by excluding metabolites that had a missingness percentage that was either below 10% or above 90% within each study. The rationale for this approach was to exclude metabolites with a high probability of having missing values due to systematic and random errors (<10%), are truly missing (>90%), or other unknown causes. Accordingly, we selected 341 and 425 metabolites in NEO and the Rhineland Study, respectively.

### Genotyping and imputation

In NEO, DNA was extracted from 6,671 venous blood samples obtained from the antecubital vein. Genotyping was performed in the Centre National de Génotypage (Evry Cedex, France), using Illumina HumanCoreExome-24 BeadChip (Illumina Inc., San Diego, California, United States of America). The detailed quality control process has been described previously.^21^ Genotypes were imputed to the Haplotype Reference Consortium (HRC) release 1.1.^22^ In the Rhineland Study, 4,165 DNA samples isolated from buffy coats extracted from blood samples were genotyped using the Illumina Omni-2.5 exome array and processed with GenomeStudio (version 2.0.5). Quality control was performed using PLINK (version 1.9). SNPs were excluded based on poor genotyping rate (< 99%) or Hardy-Weinberg Equilibrium (p < 1×10^-6^). Additionally, participants with poor quality DNA samples were excluded, on account of a poor call rate (<95%) (N=41), abnormal heterozygosity (N=69), cryptic relatedness (N=261), or a sex mismatch (N=28). To account for variation in the population structure, which may otherwise cause systematic differences in allele frequencies.^23^ We used EIGENSTRAT (version 16000). EIGENSTRAT uses principal component analysis to detect and correct for population structure, which resulted in the exclusion of an additional 164 participants from non-European descent. Finally, imputation was performed with IMPUTE (version 2),^24^ using the 1000 Genomes version 3 phase 5 as the reference panel.^25^

### Genome-wide Association Analyses

In NEO, we performed the GWAS of missing metabolites using the SNPTEST v2 software, employing logistic regression analysis under an additive model. In the Rhineland Study, the GWAS was performed using the REGENIE software (v2.2),^26^ fitting a firth logistic regression model to the data. REGENIE computation is composed of two steps. Step 1 uses a subset of genetic markers to fit a whole genome regression model that captures the phenotype variance attributable to genetic effects. In step 2, a larger set of imputed SNPs are used in order to test for their association with the different phenotypes conditional upon the prediction from step 1 and using a leave one chromosome out scheme. Genotyped SNPs were pruned using a linkage disequilibrium (LD) r²-threshold of 0.9 with a window size of 1,000 and a step size of 100 markers.

Overall, we included 21,243,072 and 49,953,404 imputed SNPs in the GWAS analysis in NEO and the Rhineland Study, respectively. These analyses were restricted to SNPs with an imputation quality > 0.3 and minor allele frequency (MAF) > 0.01. The missing metabolites were adjusted for age, sex, fasting status and five genetic principal components in the NEO study, and age, sex, fasting status and the first ten genetic principal components in the Rhineland Study. Genome-wide significance level was set at p < 5×10^-8^. However, because of the large number of outcome variables (i.e., missing metabolites), we corrected for multiple testing using the method of Li & Ji,^27^ which estimates the effective number of independent tests. Accordingly, we estimated the effective number of independent tests to be 315 and 313 in NEO and the Rhineland Study, resulting in a metabolome-wide significance level of p < 1.58×10^-10^ (≍ 5×10^-8^/315) and p < 1.59×10^-10^ (≍ 5×10^-8^/313), respectively.

### Meta-analysis of GWAS

For the GWAS meta-analysis, we selected and used 7,310,783 SNPs which had MAF > 0.01 in the Rhineland Study, as it had the larger sample size between the two studies. We then harmonized the SNPs with the overlapping 6,610,552/7,310,783 SNPs in the NEO study that had a MAF > 0.01. Meta-analysis was performed employing an inverse variance-weighted fixed-effects model using METAL.^28^ Identification of allele flips and applying genomic control was performed using METAL as well for each cohort prior to performing the meta-analysis. The metabolome-wide significance level for the meta-analysis was set at p < 1.59×10^-10^, which was the more stringent cut-off used in the Rhineland Study.

### Definition of Genomic Risk Loci

To identify genomic regions associated with missing metabolites, a single dataset was created by identifying the minimum p-value for each SNP across all meta-GWAS summary statistics of missing metabolites (6). This dataset was LD-clumped (r2 < 0.6) using the Functional Mapping and Annotation (FUMA) platform^29^ with the 1000 Genomes Phase 3 European reference panel to account for the LD structure. We further represented those clumped SNPs by lead SNPs, which are a subset of the independent significant SNPs that are in approximate LD with each other at r^2^ > 0.1. Finally, we identified associated genomic risk loci by merging any physically overlapping lead SNPs (LD blocks < 250 kb apart).

### Novel Associations

To identify novel GWAS hits, we used curated metabolome GWAS databases such as mGWAS-Explorer^30^ and PhenoScanner.^31^ We further manually verified whether our lead SNPs were previously identified in metabolomic-based GWAS studies^10^ as metabolome quantitative trait locus (mQTLs).

### Identifying Candidate Genes

We aimed to identify candidate genes tagged by the lead SNPs that may influence the probability of missingness of certain metabolites. To achieve this, we first used the “prioritization of candidate causal genes at molecular QTLs” (ProGeM) framework.^32^ This framework implements a two-step approach (bottom-up and top-down) to identify putative causal genes. In the bottom-up approach, the three closest protein-coding genes within 500 kb of the lead SNP are selected, while the top-down approach uses curated gene function databases (e.g., Gene Ontology (GO), Kyoto Encyclopedia of Genes and Genomes (KEGG), Mouse Genome Informatics (MGI) and Orphanet) to identify biologically relevant genes that are present within 500 kb of the lead SNPs. We used Variant Effect Predictor (VEP)^33^ to search for the closest protein coding genes with the lead SNP and also calculated the impact factor score of the lead SNPs based on its function as either missense, start loss or stop gain. ProGEM also assesses whether the lead SNPs are eQTL using the GTEX v7 database.^34^ The genes that were identified through top-down and bottom-up approach, were prioritized as candidate genes.

### PheWAS

We performed a phenome-wide association study (PheWAS) using the disgenet2r package to check which lead SNPs had previously been reported to be associated with any other clinical or disease outcomes, as contained in the disease-gene association database (DisGeNET). The package ranks the associations using Variant Disease Association (VDA) scores ranging from 0 to 1, where a higher score represents stronger evidence of a SNP association with a disease outcome. Only lead SNPs with a score > 0.7 were reported.

### RNA sequencing data in the Rhineland Study

Total RNA was isolated from 3,384 whole blood samples, stored, and stabilized in PAXgene Blood RNA tubes (PreAnalytix/Qiagen) using PAXgene Blood miRNA Kit in accordance with the manufacturer’s instructions and following the semi-automatic purification protocol (PreAnalytix/Qiagen). RNA integrity and quantity were assessed using the Tapestation 4200 system (Agilent). After using 750 ng of total RNA to generate next generation-sequencing libraries for total RNA sequencing (TruSeq stranded total RNA kit,Illumina), a Ribo-Zero Globin reduction was performed. Libraries were quantified using Qubit HS dsDNA assay (Invitrogen) and clustered at 250 pM concentrations on a NovaSeq6000 instrument using NovaSeq S2 v1 chemistry (Illumina) in XP mode for the first batch of 3,000 samples and NovaSeq S4 v1.5 chemistry for the last batch of 384 samples and sequenced paired-end 2*50 cycles. Sequencing data was demultiplexed and converted into fastq format using bcl2fastq2 v2.20. We performed the quality control check on raw sequencing reads using FastQC v0.11.9 and we filtered low-quality score reads using Trimmomatic v.0.39. Next, we used STAR v2.7.1 aligner to align the sequencing reads to the human reference genome GRCh38.p13 and to generate the gene count matrix through “STAR –quantMode GeneCounts” using the human gene annotation version GRCh38.101. Genes with overall mean expression greater than 15 reads and expressed in at least 5% of the participants were selected for further analysis. Finally, we used the “varianceStabilizingTransformation” function from DESeq2 v1.30.1 to normalize and transform the raw counts.

### Gene Expression quantitative trait loci

To functionally validate the GWAS results using gene expression data, we used a three-step approach analysis conducted on the first 3,384 consecutive participants of the Rhineland Study on which genetics, gene expression and metabolite data were available and quality controlled (N = 2,575). First, we assessed the associations between the lead SNPs and the corresponding genes, selected through bottom-up and top-down approaches. We adjusted gene expression levels for age, sex, the first 10 principal components, red and white blood cell counts, the relative fractions of basophils, eosinophils, lymphocytes, monocytes and neutrophils, and batch effect and we extracted the residuals. Next, linear regression analysis was performed to assess the associations between the lead SNPs (independent variable) and the residuals of candidate genes (dependent variable). Second, we evaluated the relations between the residuals of gene expression data, obtained after adjustment for identical covariates as before (independent variable) and the significant metabolites with missing values, adjusting for metabolomics’ batch effect using logistic regression analysis. Third, we employed a mediation analysis using the R package lavaan v.06-11 to investigate which candidate genes mediated the associations between lead SNPs and missing metabolite with 1000 bootstrapping iterations.

### Protein quantitative trait analysis

We performed a protein quantitative trait analysis by using the Phenoscanner. ^31^ Briefly, Phenoscanner holds publicly available protein quantitative trait loci (pQTLs) results from large-scale genome-wide association studies. We verified whether our lead SNPs associated with missing metabolites were previously identified as pQTLs. Accordingly, we filtered results for protein wide association studies with significant SNP-protein associations (p<5×10^-8^) with our lead SNP or a proxy SNP (r2>0.9) with our lead SNPs.

### Network Representation of Gene-SNP-metabolite Associations

We used the associations identified through our meta-analysis, ProGEM mapping, and PheWAS to construct a comprehensive interactive network. Each SNP, metabolite, gene, metabolite sub-pathway (as annotated in the metabolon dataset), disease or phenotype associations from DisGeNET, and pQTL associations from Phenoscanner were presented as “nodes” with distinct colours. The width of the “edges” connecting the SNP-metabolites was determined according to the -log10 of the p-value of the effect estimate. Additionally, the colour of these edges was chosen to reflect the novelty of the association. Visualization and layout of the networks were created using Cytoscape version 3.10.2 and Gephi v0.10 and then exported as an interactive HTML5 using the sigmaExporter plugin ^35^ at the following URL https://tofaquih.github.io/GWASMissingMetabolites/.

## Results

### Discovery Genome-wide Association Studies of Missing Metabolites

The GWAS of missing metabolite measures was performed separately in 594 individuals from the NEO study (mean (standard deviation (SD)) age: 55.8 (5.9), range: 45-66 years, 53% women), and 4,165 individuals from the Rhineland Study (mean (SD) age: 55.5 (14), range: 30-96 years, 56% women). Individual study characteristics and general genotype assay information are summarized in **Table 1**. GWAS results in NEO identified 712 metabolome-wide significant (p<1.58×10^-10^) associations between 537 SNPs and 6 out of the 341 included metabolites. In the Rhineland Study, we identified 4,370 metabolome-wide significant (p<1.59×10^-10^) associations between 2,615 SNPs and 32 out of the 425 included metabolites. Restricted to the metabolites that were available in both studies (N=224), the study-specific GWAS identified 523 and 2,613 metabolome-wide significant SNPs for 5 and 26 metabolites in the NEO and the Rhineland study, respectively. The overall workflow of the GWAS analysis and following downstream analyses is illustrated in **Figure 1**. The summary statistics of GWAS analysis are available in **Tables S1** and **Table S2** for NEO and the Rhineland Study, respectively.

**Table 1:** Overview of the sample characteristics and general genotype assay information.

| **Characteristic** | **NEO Study**<br>**(n= 594)** | **Rhineland Study**<br>**(n= 4165)** |
| --- | --- | --- |
| Age: mean (sd) | 55.8 (4.5) | 55.5 (13.96) |
| Sex: % female | 53% | 56% |
| Metabolomics Platform | Metabolon-HD4 | Metabolon-HD4 |
| N of metabolites with missing values | 433 | 340 |
| Blood sampling | Serum | Plasma |
| Fasting status % | 100% | 99.4% |
| Genotyping Array | Illumina HumanCoreExome-24 BeadChip | Omni 2.5 Exome Array |
| Genotype Imputation Panel | HRC (r1.1) | 1000 genome phase 3 version 5 |

**Figure 1:**
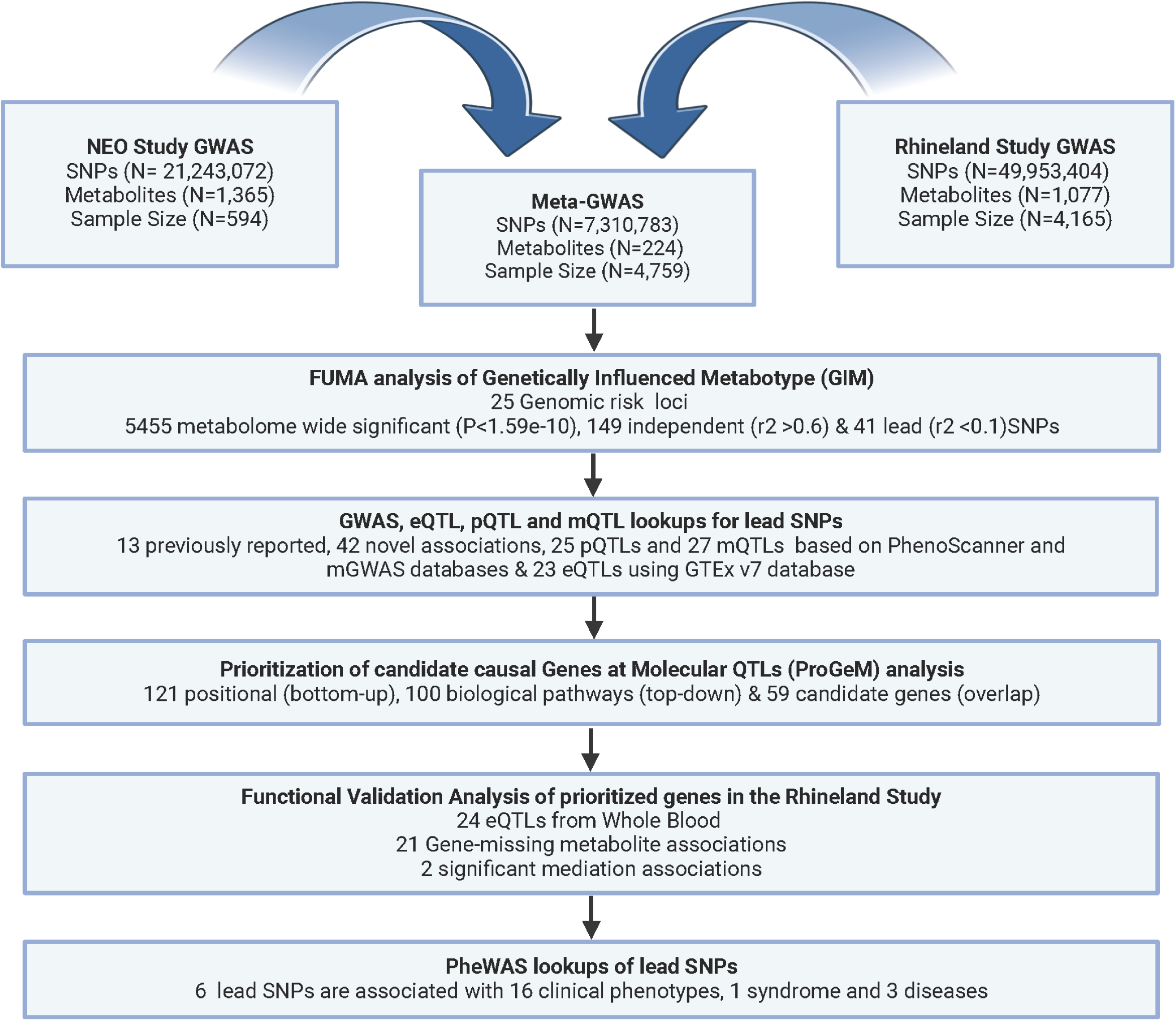
Workflow of the GWAS and post-GWAS analyses of missing metabolite measures.

### Genome-wide Meta-analysis of Missing Metabolites

A meta-analysis of the overlapping 224 metabolites in the two studies identified 5,455 significant associations (p<1.59×10^-10^), including 3,260 SNPs across 33 different metabolites (**Figure 2** and **Table S3)**. The direction of the associations was similar across both cohorts (Pearson correlation R2=0.92) using independent SNP-metabolite (r^2^<0.6) associations (**Figure S1** and **Table S4**). The majority of these metabolites belonged to the steroid metabolism pathway (N=7), followed by amino-acid metabolism (N=4), fatty acid metabolism (N=4), bile acid metabolism (N=5), and unannotated metabolites (N=8). Other hits belonged to food and plant-derived xenobiotics (i.e., alliin, solanidine, ferulic acid 4-sulfate and caffeic acid sulfate) and nucleotide metabolites (xanthosine).

**Figure 2:**
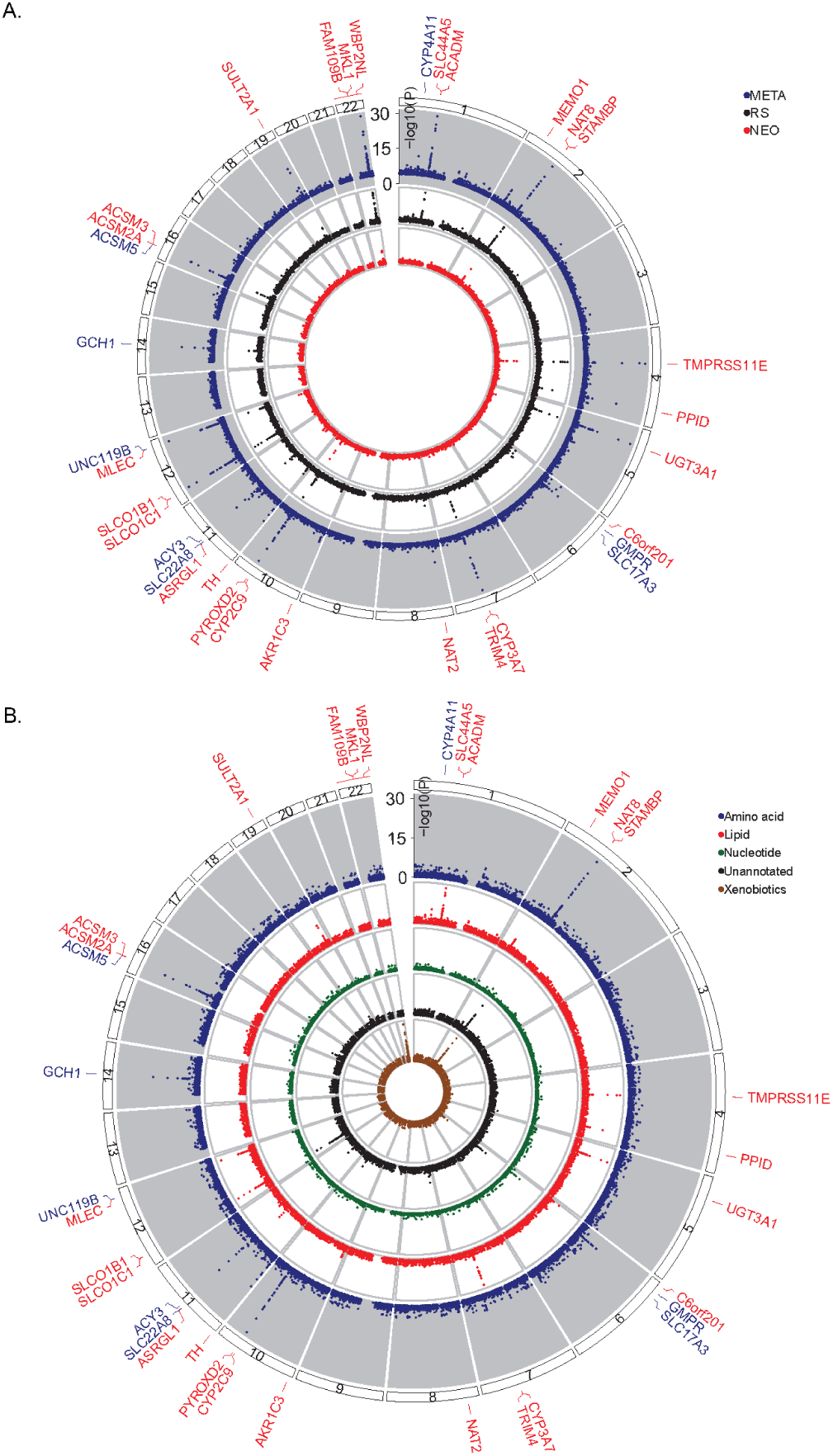
Genomic loci associated with missingness of metabolites. **A,** Circular Manhattan plot showing both cohort-specific and meta-analysis GWAS results. **B,** Circular Manhattan plot showing regional associations of genomic locations per metabolite class. The color of the genes indicates whether the identified lead SNP locus is novel. Dark blue indicates lead SNP loci (r2<0.1) that were previously reported, and red lead SNP loci that were identified to be novel with metabolites. The p-value axis is truncated at p<1×10^-30^ for visualization purposes. All GWAS models were adjusted for age, sex, batch, fasting status and population substructure principal components.

### Genetically Influenced Metabotypes

Genetically Influenced Metabotypes (GIMs), defined as variant-metabolite clusters^7^, were identified by merging the summary statistics of all SNP-metabolite associations and selecting the SNPs with the lowest association P-value, resulting in a set of 7,310,783 unique SNPs. Functional Mapping and Annotation (FUMA) identified 3,260 metabolome-wide significant SNPs indexed by 41 lead SNPs (linkage disequilibrium (LD) r^2^ < 0.1) located across 25 genomic risk loci (**Table S4** and **Table S5**). Those 41 lead SNPs had a total of 55 associations with the odds of missingness of 28 metabolites. Those 55 lead SNP-metabolite associations were further cross-referenced with previous metabolomic GWAS results and metabolome-based GWAS databases to assess the novelty of the associations (**Table 2** and **Table S6**). We defined SNP-metabolite associations as “novel” if the SNP-metabolite association was not reported previously, and labelled it as “reported” otherwise. Accordingly, we found 42 novel and 13 previously reported associations. Several of the associated metabolites were connected by shared pathways and genes, as shown in **Figure 3**, and formed two large clusters. The first cluster was enriched for steroids and bile acid metabolites and contained two pleiotropic SNPs (rs4149056 and rs45446698) associated with 5 and 4 metabolites, respectively. The second cluster comprised of acetylated tryptophan and lysine related metabolites, along with the xenobiotic metabolite alliin. An interactive version of this network is available online at https://tofaquih.github.io/GWASMissingMetabolites/ for further exploration.

**Table 2:**
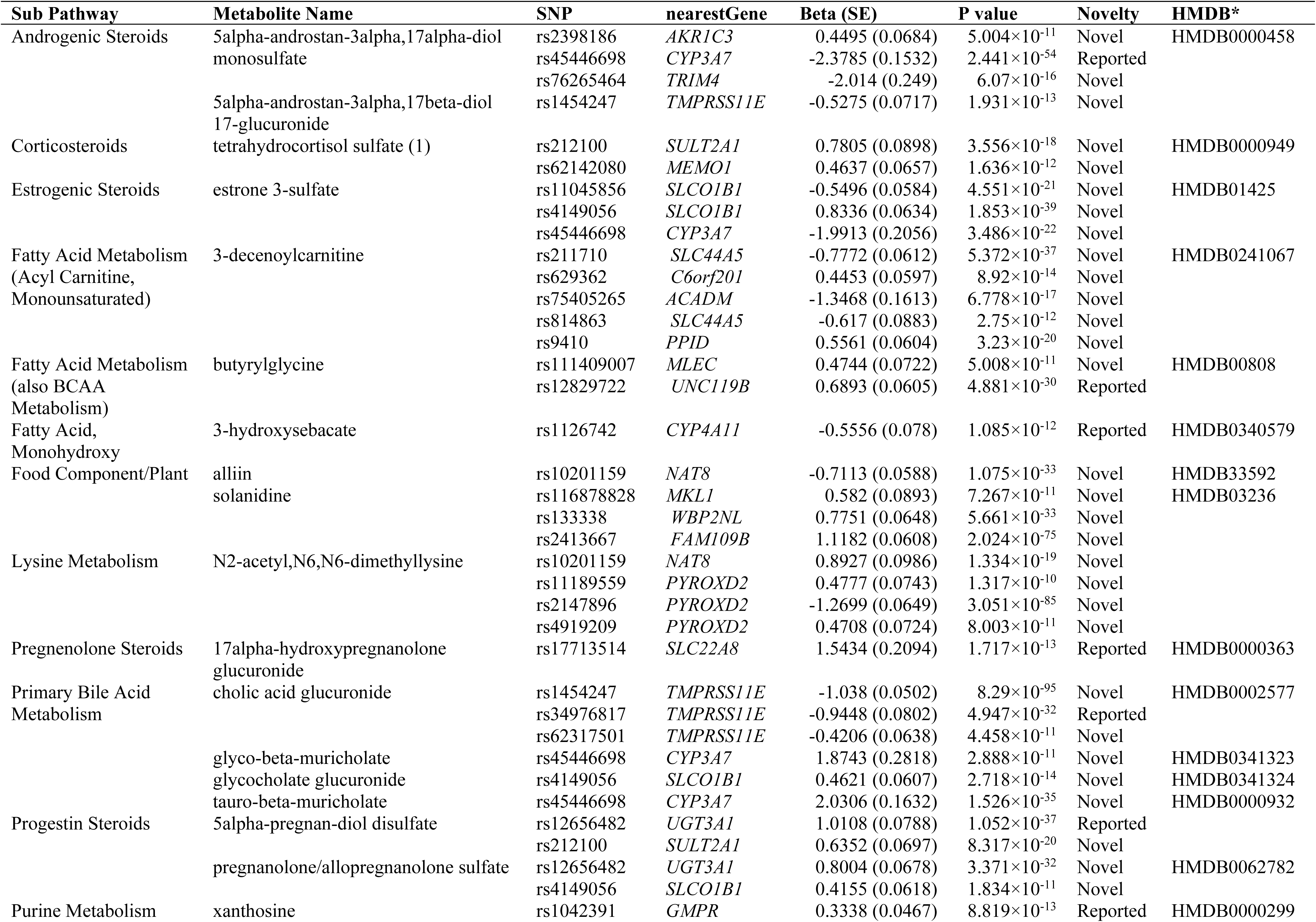
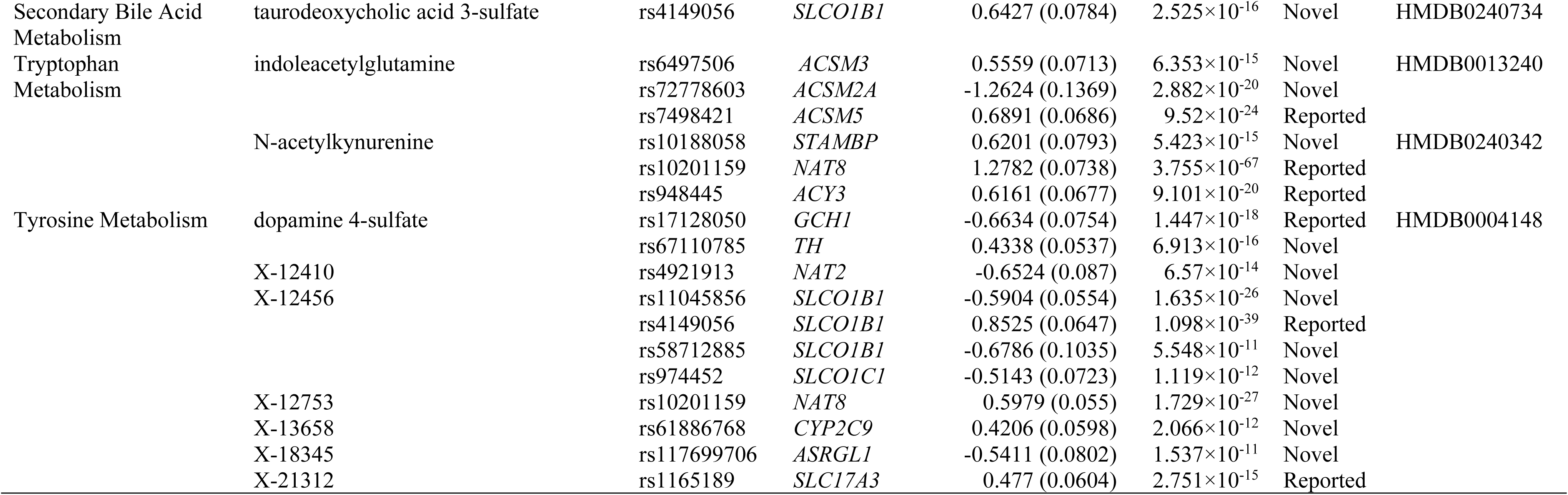
Novel and previously reported lead SNP-Metabolite associations using PhenoScanner.

**Figure 3:**
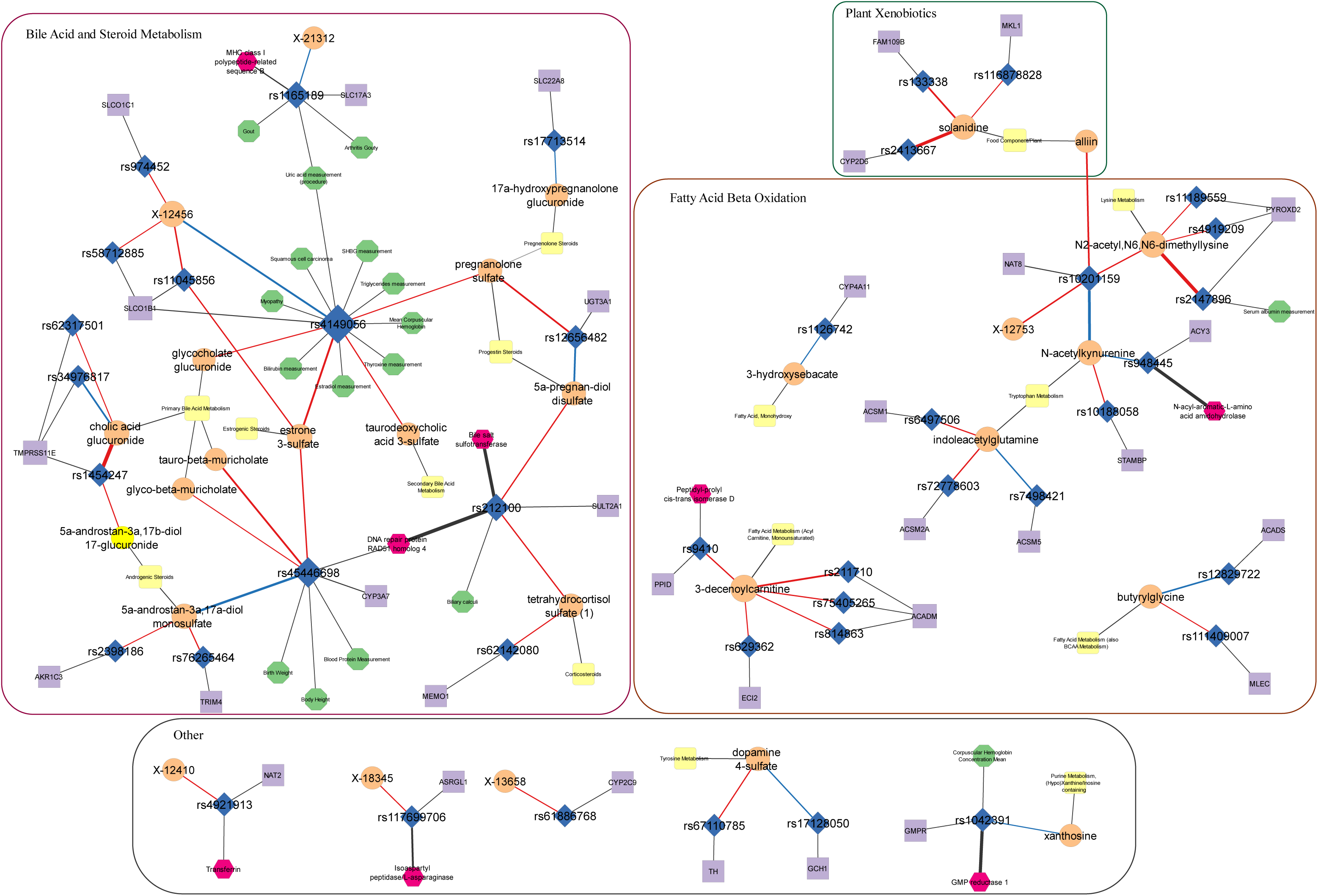
Network representation of the 41 lead SNP-metabolite associations. The network shows the associations between the 41 lead SNPs (blue diamonds) and metabolites (orange circles). The network also includes the mapped genes to each SNP (purple squares), assigned sub-pathway from the measurement platform (yellow rounded squares), traits and diseases associated with the SNPs from DisGeNET (green octagons), and pQTL associations from Phenoscanner (pink hexagons). Novel and reported associations between the SNP-metabolites are represented in red- and orange-colored lines respectively.

### Identification of eQTLs, pQTLs and mQTLs

Using GTEx (version 7), we identified 23 expression Quantitative Trait Loci (eQTLs) (**Table S6**). Interestingly, among the novel associations, the SNP rs2413667, which was associated with the odds of missingness of solanidine, was an eQTL of *CYP2D6*, an important enzyme for xenobiotic metabolism^36^, in adipose tissue. The PhenoScanner results showed that five lead SNPs were previously reported as protein Quantitative Trait Loci (pQTLs) for seven different proteins (**Table 3** and **Table S7**). These included peptidyl-prolyl cis-trans isomerase D, major histocompatibility class I polypeptide-related sequence B, and DNA repair protein RAD51 homolog. Finally, using the metabolome-based GWAS study databases, we identified 27 lead SNPs as metabolome Quantitative Trait Loci (mQTLs) (**Table S8**). For example, we identified rs211710, which was associated with the odds of missingness of 3-decenoylcarnitine in our study, as an mQTL for decenoylcarnitine.

**Table 3:** Top pQTL associations with the lead SNPs.

| Metabolite Name | Sub Pathway | Lead SNP | Nearest gene | Proxy pQTL SNPs* | pQTL associated trait (Phenoscaner) | pQTL associated trait (UK biobank) |
| --- | --- | --- | --- | --- | --- | --- |
| 3-decenoylcarnitine | Fatty Acid Metabolism (Acyl Carnitine. Monounsaturated) | rs9410 | *PPID* | rs8396 | Peptidyl-prolyl cis-trans isomerase D | - |
| X-21312 | - | rs1165189 | *SLC17A3* | rs13200784, rs3757132 | MHC class I polypeptide-related sequence B | - |
| estrone 3-sulfate | Estrogenic Steroids | rs45446698 | *CYP3A7* | rs45446698 | DNA repair protein RAD51 homolog 4 | - |
| tauro-beta-muricholate | Primary Bile Acid Metabolism |  |  |  |  | - |
| 5a-Androstane-3a,17a-diol monosulfate | Androgenic Steroids |  |  |  |  | - |
| glyco-beta-muricholate | Primary Bile Acid Metabolism |  |  |  |  | - |
| X-12410 | - | rs4921913 | *NAT2* | rs4921915 | Transferrin | - |
| 5a-Androstane-3a,17a-diol disulfate | Progestin Steroids | rs212100 | *SULT2A1* | rs212100 | Bile salt sulfotransferase | SULT2A1 |
| tetrahydrocortisol sulfate | Corticosteroids |  |  |  | DNA repair protein RAD51 homolog 4 |  |
| X-18345 | - | rs117699706 | *ASRGL1* | - | - | Isoaspartyl peptidase/L-asparaginase |
| N-acetylkynurenine (2) | Amino acid | rs948445 | *ACY3* | - | - | N-acyl-aromatic-L-amino acid amidohydrolase (carboxylate-forming) |
| xanthosine | Nucleotide | rs1042391 | *GMPR* | - | - | GMP reductase 1 |

\*Proxy SNPs selected based on their correlation ( $r^2$ ) with the lead SNP. Full results available in Supplementary Table 7.

### Prioritized Candidate Genes for Missing Metabolites

To map the 41 identified lead SNPs associated with the odds of missingness of the metabolites to candidate casual genes, we used the ‘Prioritization of candidate causal Genes at Molecular QTLs’ (ProGeM) framework. This framework maps SNPs to genes using two complementary methods. The first is based on positional proximity, which mapped the identified SNPs to 121 genes. The second is based on biological relevance, which mapped the SNPs to 100 relevant genes (**Figure 4**, **Table S9** and **Table S10**). Subsequently, we focused the analysis on genes that were mapped through both methods, resulting in 59 candidate causal genes.

**Figure 4:**
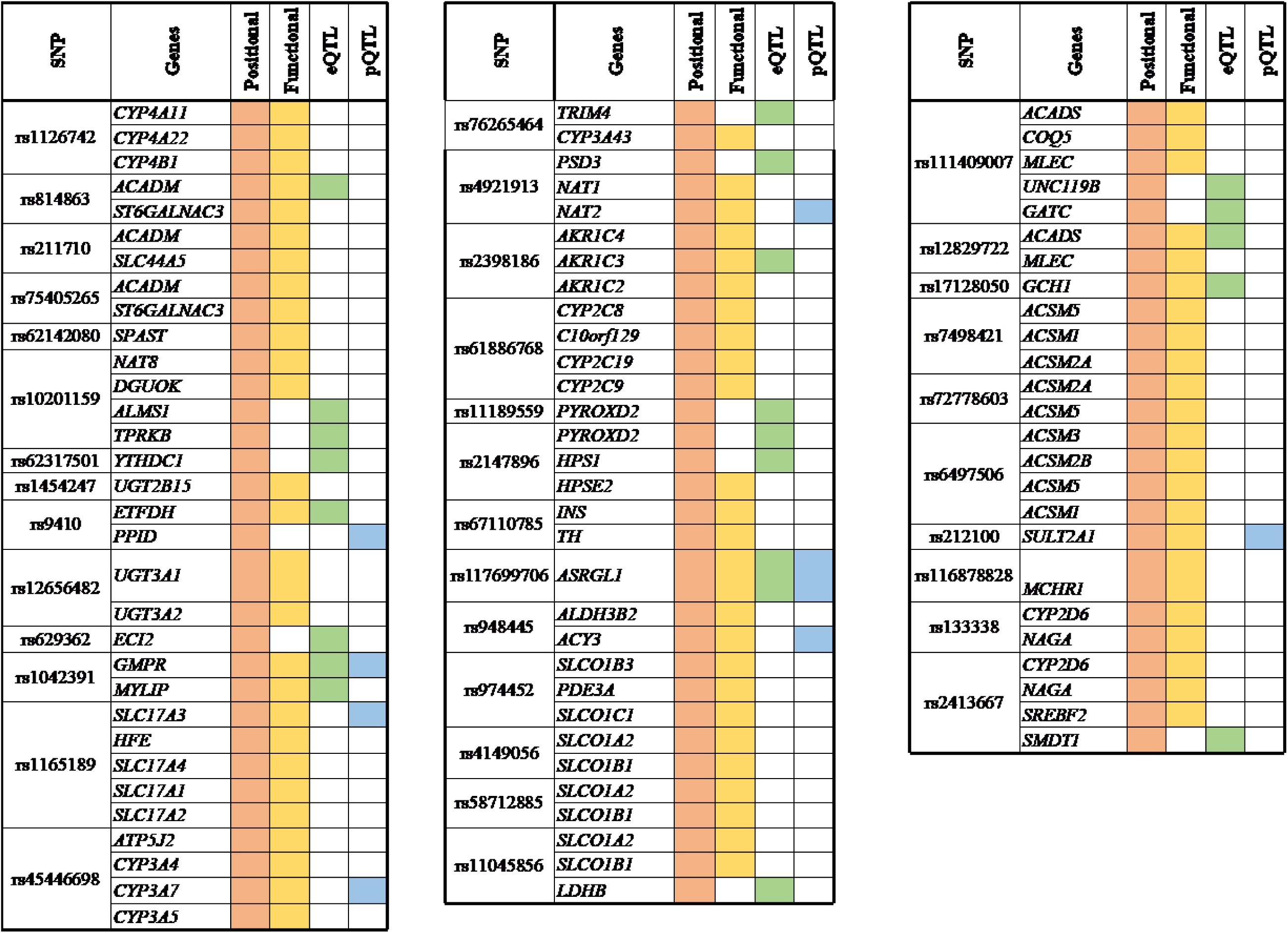
Summary of top ranked candidate gene identification, eQTL, and pQTL associations per identified SNP-metabolite. Candidate genes identified by the ProGeM positional approach (bottom-up) are highlighted in orange under the “positional” column and highlighted in yellow under the “functional” column based on metabolic and phenotypic relevance approach (top-down). Genes with a significant eQTL association are highlighted in green under the eQTL column and in blue for SNP-pQTL associations under the pQTL column.

### Mediation Analysis Between SNP, Gene Expression Levels, And Odds of Missingness

Out of the 162 identified genes, we conducted a functional analysis for 78 genes whose expression levels were available in 2,575 participants of the Rhineland Study. We found that 18 lead SNPs were significantly associated with 20 genes at a nominal p-value threshold—with the relation between rs10201159 and *ALMS1* emerging as the strongest association (β = -1.55, p-value < 0.001) (**Table S11)**. The expression levels of 18 genes were significantly associated with the odds of missingness for 11 metabolites—with the *HPS1* and N2-acetyl, N6,N6-dimethyllysine association emerging as the strongest (β = -0.4532, p-value < 0.001). Finally, mediation analysis (**Table 4**) conducted involving 9 genes whose expression levels were significantly associated with both lead SNPs and the corresponding metabolites, indicated that *SMDT1* and *HPS1* partially mediated the relation between the SNPs and the odds of missingness of the corresponding metabolites (p<0.05). *SMDT1* mediated the effect between rs2413667 and solanidine by 3.2% (β (SE) = -0.02 (0.01)), while *HPS1* mediated the effect between rs2147896 and N2-acetyl, N6,N6-dimethyllysine by 2.8% (β (SE) = -0.02 (0.01)).

**Table 4:** eQTL mediation of the association between the lead SNPs and missing metabolites by primary candidate genes.

| Exposure | Mediator | Metabolite | Indirect effect |  | Direct effect |  | Total effect |  | Percentage of mediation<sup>±</sup> |
| --- | --- | --- | --- | --- | --- | --- | --- | --- | --- |
|  |  |  | β estimate (SE) | P value | β estimate (SE) | P value | β estimate (SE) | P value |  |
| rs117699706 | *ASRGL1* | X-18345 | -0.030<br>(0.023) | 0.212 | -0.30<br>(0.067) | 6.6×10<sup>-06</sup> | -0.333<br>(0.062) | 9.6×10<sup>-08</sup> | 8.9% |
| rs629362 | *ECI2* | 3-Decenoylcarnitine | -0.015<br>(0.013) | 0.259 | -0.236<br>(0.047) | 7.6×10<sup>-07</sup> | -0.252<br>(0.045) | 2.9×10<sup>-08</sup> | 6.2% |
| rs10201159 | *ALMS1* | X-12753 | -0.025<br>(0.025) | 0.320 | -0.407<br>(0.053) | 1.8×10<sup>-14</sup> | -0.433<br>(0.046) | <10×10<sup>-16</sup> | 5.9% |
| rs814863 | *ACADM* | 3-Decenoylcarnitine | -0.011<br>(0.007) | 0.172 | -0.319<br>(0.063) | 4.9×10<sup>-07</sup> | -0.330<br>(0.063) | 2.0×10<sup>-07</sup> | 3.2% |
| rs2413667 | *SMDT1* | Solanidine | -0.024<br>(0.012) | 0.055* | -0.722<br>(0.050) | <10×10<sup>-16</sup> | -0.746<br>(0.047) | <10×10<sup>-16</sup> | 3.2% |
| rs10201159_T | *ALMS1* | N2-acetyl.N6.N6-dimethyllysine | -0.015<br>(0.035) | 0.684 | -0.495<br>(0.074) | 2.9×10<sup>-11</sup> | -0.509<br>(0.066) | 1.3×10<sup>-14</sup> | 2.9% |
| rs2147896 | *HPS1* | N2-acetyl.N6.N6-dimethyllysine | -0.022<br>(0.007) | 0.004* | -0.745<br>(0.046) | <10×10<sup>-16</sup> | -0.766<br>(0.046) | <10×10<sup>-16</sup> | 2.8% |
| rs11189559_G | *PYROXD2* | N2-acetyl.N6.N6-dimethyllysine | -0.005<br>(0.004) | 0.176 | -0.236<br>(0.049) | 1.9×10<sup>-06</sup> | -0.241<br>(0.049) | 1.1×10<sup>-06</sup> | 2.2% |
| rs211710 | *ACADM* | 3-Decenoylcarnitine | -0.002<br>(0.006) | 0.731 | -0.556<br>(0.048) | <10×10<sup>-16</sup> | -0.557<br>(0.048) | <10×10<sup>-16</sup> | 0.4% |

\*Indirect effect's P value =< 0.05
<sup>±</sup>(Indirect effect / total effect) \* 100

### Missing Metabolites Variants Implicated in Diseases

Our search in the DisGeNET database, using curated data, revealed that six lead SNPs were related to sixteen phenotypes, three diseases and one syndrome (**Table S12**). The phenotypes were either related to liver, kidney, or reproductive function. Notably, rs4149056 was associated with 7 phenotypes including levels of thyroxine, sex hormone binding globulin (SHBG), and estradiol. Another notable SNP was rs45446698, which was related to three phenotypes, including birth body weight, body height, and blood protein measurement. Lastly, in the disease category, we found rs4149056 SNP to be associated with squamous cell carcinoma.

## Discussion

We conducted a genome-wide meta-analysis to identify genetic variants associated with the odds of missingness of metabolites and identified 55 lead SNP-‘missing metabolite’ associations, of which 42 associations were novel (i.e., not found in previous GWAS of metabolite levels). Based on comprehensive *in silico* functional analyses, we identified associations between specific groups of metabolites and common metabolic pathways. First, we identified several SNP-metabolite associations involving metabolites that play a biological role in fatty acid beta oxidation pathways—generally containing an acetyl group—and that are also expressed or related to kidney function. Second, we found a group of metabolites and SNPs related to bile acid and steroid metabolism. And third, we identified SNPs associated with two xenobiotics primarily derived from plant sources.

Regarding the first group of SNP-metabolites associated with fatty acid beta oxidation, notable findings were related to the following metabolites: 1) 3-decenoylcarnitine, a medium-chain acylcarnitine that is involved in the production of energy via beta-oxidation by transporting acyl-groups into mitochondria^37^; we found five novel associations between intronic regions of *ACADM* and *ECI2,* as well as exonic loci in *PPID (*also known as *CypD* ^38^) and 3-decenoylcarnitine’s odds of missingness, 2) indoleacetylglutamine, a gut microbiome–derived metabolite^39^ involved in tryptophan metabolism and has been reported in various studies regarding gut microbiome and chronic kidney disease.^40,41^ Two novel associations near the *ACSM1* and *ACSM2A* genes were associated with its odds of missingness, 3) N-acetylkynurenine, another metabolite involved in tryptophan metabolism. A novel association in the *STAMPB* gene and a novel association (rs10201159) in the intergenic region of *NAT8* were associated with its odds of missingness, 4) N2-acetyl,N6,N6-dimethyllysine, an amino acid and is the precursor of N6,N6,N6-trimethyl-L-lysine (also known as trimethyllysine (TML)), which in turn has been reported as a potential precursor for trimethylamine and trimethylamine N-oxide.^42,43^ The rs10201159 near *NAT8* and three novel associations near *PYROXD2* gene were associated with its odds of missingness.

All five novel loci associated with 3-decenoylcarnitine were located in the vicinity of genes that were strongly linked to the regulation of fatty acid beta-oxidation.^44–46^ In addition, disruptions of the metabolism of acylcarnitine and the process of beta-oxidation are well known causes for specific IEM. *ACADM* in particular is linked to medium chain acyl-CoA dehydrogenase (MCAD) deficiency,^45,47^ supporting our hypothesis of genetic variations affecting the missingness of 3-decenoylcarnitine. The pQTL analysis showed that rs9410 had been associated with PPID protein expression. Since the protein coded by *PPID* (CypD) functions as a transport pore in the mitochondrial membrane,^48^ this mutation could affect the transport of 3-decenoylcarnitine in the mitochondria and lead to reduction of its levels below the limit of detection. In addition, our functional validation analyses using gene expression data showed that *ACADM* and *ECI2* expression was associated with their respective genetic variants, as well as with those of 3-decenoylcarnitine. However, the upstream metabolite carnitine had no missingness in our data (none in NEO and only missing in 9 individuals in the Rhineland study). Additionally, we observed that 3-decenoylcarnitine missingness occurred with higher carnitine levels, not when carnitine was low (**Figure S2A** and **Figure S3A**). This is an indication that these genetic variations could be affecting the metabolism or transport of carnitine to 3-decenoylcarnitine leading to an accumulation of carnitine and delayed production of 3-decenoylcarnitine. Similar to 3-decenoylcarnitine, the biological pathways implicated in indoleacetylglutamine metabolism and its respective SNP associations were related to beta-oxidation of fatty acids. Indoleacetylglutamine is also reportedly elevated in urine of patients with Hartnup disease—an IEM.^41,49^

N-acetylkynurenine is a metabolite belonging to the kynurenine pathway, which in turn are crucial for the breakdown of tryptophan. ^50^ We found the probability of N-acetylkynurenine missingness is linked to the SNP rs10188058 in the *STAMPB* gene. It is worth noting that both N-acetylkynurenine (as well as the kynurenine pathway) and *STAMPB* were previously reported to be associated with the IEM microcephaly-capillary malformation syndrome.^50,51^ N-acetylkynurenine, along with N2-acetyl,N6,N6-dimethyllysine (lysine metabolism), alliin, and X-12753, are also associated with the SNP rs10201159 in the intergenic region of *NAT8*. In line with the function of *NAT8*, this locus has been reported to be associated with several acetyl forms of amino acids and fatty acids metabolism and with the progression of chronic kidney disease.^52^ Overall, these metabolites are associated with beta-oxidation, mitochondrial function, IEMs, and potentially related to kidney function.

Although little is known about N2-acetyl,N6,N6-dimethyllysine in the literature, its precursors—namely TML— has been studied extensively.^42,43,53^ TML has also been associated with cardiovascular diseases and reportedly predicted all-cause mortality and cardiovascular disease.^43^ The loci associated with the missingness of N2-acetyl,N6,N6-dimethyllysine are near the regions of *PYROXD2* and *NAT8*. These two genes are related as *PYROXD2* has been reported to interact with *NAT8* in several studies. ^9,54,55^ *PYROXD2* is localized in the mitochondrial inner membrane and has an important role in regulating mitochondrial function.^56^ In addition, *PYROXD2* is associated with the IEM disorder trimethylaminuria.^57^ *PYROXD2* is normally associated with low levels of trimethylamine in the urine of healthy individuals.^57^ Although the commonly reported primary mutations causing this disorder are in the *FMO3* gene, growing evidence suggests that mutations in *PYROXD2* play a role as well. Indeed, genetic variations in *PYROXD2* have been reported to be implicated with the increased levels of trimethylamine in individuals with trimethylaminuria, specifically in sweat, breath, and urine.^57,58^ The eQTL analysis further supported that *PYROXD2* expression was associated with the SNPs (rs11189559 and rs2147896) and with the odds of N2-acetyl,N6,N6-dimethyllysine missingness. Our GWAS and eQTL findings could indicate the involvement of our novel SNPs associations in *PYROXD2* in relation to the missingness of N2-acetyl,N6,N6-dimethyllysine. Subsequently, these are possibly associations with poor metabolism of TML and trimethylamine that could additionally relate to the development of trimethylaminuria.

The second group of metabolites we have reported from our analysis were steroids and bile acids with a shared association with two pleiotropic SNPs: rs4149056 and rs45446698. Rs4149056 (*SLCO1B1*) was associated with two bile acids, two estrone metabolites, and an unknown metabolite X-12456—predicted to be analogous to steroid metabolites—which is in line with findings from previous genomic research literature.^10^ First, *SLCO1B1* is associated with statin-induced myopathy via the interaction with bile acids and cholesterol.^59^ Second, rs4149056 and *SLCO1B1* were reported to be associated with serum estrone levels.^60,61^ Similarly, rs45446698 (*CYP3A7*) has been reported in studies relating breast cancer to oestrone and progesterone levels,^62,63^ as well as studies related to atorvastatin metabolism.^64^ In addition, PheWAS analysis indicated previous associations between rs45446698 and birth weight, a trait with lifetime implications for metabolism.^65^ Overall, these two SNPs participate in similar metabolic processes affecting steroid and bile acids. Although it remains unclear how these two SNPs are involved in reducing the metabolite levels to missingness levels, our pQTL suggests that rs45446698 is associated with post-translational modification of DNA repair protein RAD51 homolog 4 (RA51D) in a trans manner.^66,67^ These modifications could indicate consequences of the rs45446698 SNP on the RAD51 functionality that could contribute to the missingness of estrone 3-sulfate, tauro-beta-muricholate, “5alpha-androstan-3alpha,17alpha-diol monosulfate”, and glyco-beta-muricholate.

The final group we have identified was comprised of alliin and solanidine, which are both xenobiotic metabolites derived from the consumption of garlic and potatoes, respectively. Alliin is generally known for its health benefits, such as improved glucose tolerance in mice and anti-inflammatory effects in rats and *in vitro* studies.^68,69^ Interestingly, the *NAT8* locus was found to be associated with the odds of missingness of alliin. A previous study reported another SNP in *NAT8* to be associated with the acylated form of alliin—N-acetylalliin.^10^ These findings indicate that *NAT8* engages in the metabolism and acylation of alliin in some capacity. It is possible that rs10201159 leads to a faster metabolism of alliin into N-acetylalliin. Consequently, the levels of alliin could fall below the detection limit and are therefore reported as missing in metabolomic analyses.

Solanidine is a steroidal alkaloid, slightly toxic metabolite in low quantities derived from potatoes and other plants of the Solanaceae family.^70,71^ We identified three novel SNPs associations from three genes strongly associated with solanidine. The rs2413667 SNP in the eQTL region of *CYP2D6* was particularly noteworthy. The coded protein from this gene is responsible for the metabolism of approximately 25% of drugs used in clinical settings and its association with solanidine has been studied in relation to metabolism efficiency.^72,73^ Solanidine has also been reported as a potential dietary marker to assess the efficiency of *CYP2D6* functionality.^74^ This was further supported by a clinical trial reporting *CYP2D6* inhibition to be associated with up to 4.56 fold increase of solanidine levels, indicating compromised xenobiotic metabolism. Therefore, additional studies used solanidine as a biomarker to identify “poor metabolizers”. ^72,74,75^ Based on our findings regarding the association of rs2413667 and the odds of missingness of solanidine in tandem with previous studies examining solanidine and *CYP2D6* metabolism, rs2413667 may be utilized as a new pharmacogenomic marker to identify poor metabolizers (or conversely rapid metabolizers) of drugs ^76^ and could be utilized in identifying and developing personalized nutritional interventions. ^77^ This may also be true for all the reported SNPs and metabolites in this study. The rs2413667 SNP is also in proximity of the eQTL region of *SMDT1* and based on our mediation analysis, this eQTL has a significant mediation effect of 3.2%. The *SMDT1* encoded protein from this gene partakes in forming a calcium uniporter complex in the mitochondria. In line with the protein function, solanidine and solanine toxicity are characterized by the disruption of calcium transport in mitochondria.^71^ Therefore, rs2413667 may affect solanidine metabolism through its influence on *SMDT1* expression.

Four pleiotropy patterns characterized the total 55 associations (**Figure 3**). First, the odds of missingness of 13 metabolites was associated with multiple loci in different genes, as was the case, e.g., for 3-decenoylcarnitine. Second, five loci illustrated pleiotropy and were associated with the odds of missingness of multiple metabolites, usually belonging to similar metabolic pathways. A noteworthy case of this was rs4149056, which was associated with the odds of missingness of five metabolite measures—three of which were steroid metabolites. Third, we observed three instances where multiple SNPs within the same gene were associated with the odds of missingness of corresponding metabolites. For example, three SNPs in *PYROXD2* were associated with the odds of missingness of N2-acetyl,N6,N6-dimethyllysine. Fourth, the remaining associations were exclusively single SNP-metabolite associations. Taken together, our findings indicate considerable genetic pleiotropy regarding the odds of missing metabolite measures, which, however, converge on common metabolic pathways.

An important consideration for this study is the interpretation of results originating from a non-traditional phenotype—missingness of metabolites. Missingness of a metabolite measurement can be caused by either a technical issue, i.e. failure of metabolite identification in the spectral data due to a deconvolution issue, or the metabolite concentration being below the limit of detection, or real missingness, i.e. the metabolite concentration is null. It can be expected that failure of metabolite detection in the spectral data due to a deconvolution issue is to an extent random and unlikely to be caused by genetics. However, it is extremely difficult to distinguish between a metabolite measure being below the limit of detection and a metabolite truly being absent, such as the case of dopamine 4-sulphate. The SNP rs67110785 is found to be associated with missingness of dopamine 4-sulphate measures and is located in close proximity to the tyrosine hydroxylase (TH) encoding gene—a rate limiting enzyme in the synthesis of dopamine.^78^ The same SNP is also an eQTL for *TH* in the Genotype-Tissue Expression (GTEx) database. At face value, TH deficiency would lead to missingness of dopamine 4-sulphate; however, TH deficiency also leads to severe neurological problems that were not reported by any of the participants. Dopamine 4-sulphate is produced from dopamine by the enzyme sulfotransferase family 1A member 3 (SULT1A3), which also produces dopamine 3-sulphate.^79^ When plotting the levels of dopamine 4-sulphate against dopamine 3-sulphate in the NEO study, it was clear that the individuals with missing values of dopamine 4-sulphate had low levels of dopamine 3-sulphate and could thus not be TH or SULT1A3 deficiency (**Figure S2B** and **Figure S3B**). Missingness of dopamine 4-sulphate is therefore likely due to the measures being below the limit of detection, probably caused by lower levels of dopamine, rather than its absence.

Although the exact mechanism through which these SNPs would induce missingness of metabolites remains to be fully elucidated, findings from previous studies related to IEM and poor metabolism, as well as our eQTL and pQTL analyses support the hypothesis that genetic factors influence the probability of metabolites absence. Future studies are needed for deeper investigation of the underlying biological pathway of the missingness of the reported metabolites. A limitation of our study is the relatively small samples sizes used in the GWAS. However, by using two studies and a meta-analysis approach, we found strong associations between the genetic variants and missingness, despite the sample size limitation, and were able to replicate our findings. A second potential limitation was using different blood sample collection methods, with serum used in NEO and plasma in the Rhineland study. The differing blood sampling methodology could have influenced the levels and detection of some metabolites and may explain some of the disparity in the total measured metabolites between the two studies. However, we did find a very strong correlation between the loci-metabolites effect estimates from the NEO and Rhineland study. Additionally, high correlations were previously reported for metabolite measurements from serum and plasma samples collected from the same individuals.^80^ Therefore, the choice of blood sampling type may have a limited impact on the overall metabolite profiles and our findings. A third limitation was the nature of the untargeted platforms. These platforms can be prone to missing data due to systematic errors.^12,13^ We accounted for this limitation by excluding metabolites outside the missingness limits (<10% or >90% missingness) to avoid the inclusion of metabolites that were simply missing due to systematic errors as much as possible. Future targeted metabolomics studies that measure the absolute concentrations of our reported metabolites can aid in replicating our findings. Finally, our study included populations from European ancestry only and, therefore, further studies are required to investigate metabolite missingness in different populations and ethnicities.

In summary, we identified 55 associations between genetic variants and the odds of missingness of numerous metabolites, 42 of which were completely novel associations. These associations involved 24 SNP-metabolite pairs related to fatty acid beta oxidation and kidney function. In addition, two pleiotropic SNPs were notable for their associations with metabolites partaking in steroid and bile acid metabolism, as well as metabolism of dietary and xenobiotic metabolites. Our results provide novel insights into the role of genetics in determining the absence of certain metabolites, with potential implications for the identification of both “poor metabolizers” and IEM. Indeed, the novel genetic variants reported here could have potential value in future etiological and prediction studies, especially in the fields of metabolomics, nutritional epidemiology, pharmacogenomics, and IEM disorders.

## Author Contributions

T.O. Faquih: Conceptualization, Methodology, Formal Analysis, Writing – Original Draft Preparation, Visualization; M.A. Imtiaz: Conceptualization, Methodology, Formal Analysis, Writing – Original Draft Preparation, Visualization; V. Talevi: Methodology, Formal Analysis, Writing – Reviewing and Editing; E.N. Landstra.: Conceptualization, Methodology, Writing – Reviewing and Editing; A. van Hylckama-Vlieg: Conceptualization, Supervision, Writing – Reviewing and Editing; N. A. Aziz: Conceptualization, Methodology, Writing – Review and Editing, Supervision; R. Li-Gao: Writing – Review and Editing; F.R. Rosendaal.: Study design, Funding acquisition, Conceptualization; R. Noordam and D. van Heemst: Funding acquisition, Writing – Review and Editing; D.O. Mook-Kanamori: Conceptualization, Methodology, Resources, Writing – Reviewing and Editing, Funding Acquisition, Supervision; M.M.B. Breteler: Conceptualization, Methodology, Resources, Writing – Reviewing and Editing, Data Curation, Funding Acquisition, Supervision; K.Willems van Dijk: Conceptualization, Supervision, Writing – Reviewing and Editing. All authors read and approved the final manuscript.

## Acknowledgements

We would like to thank all participants and the study personnel of the Rhineland Study. The authors of the NEO study thank all participants, all participating general practitioners for inviting eligible participants, all research nurses for data collection, and the NEO study group: Pat van Beelen, Petra Noordijk, and Ingeborg de Jonge for study coordination, laboratory, and data management.

## Ethical Approval

The NEO study was approved by the medical ethical committee of the Leiden University Medical Centre (LUMC) and all participants gave their written informed consent. The ethics committee of the medical faculty of the University of Bonn approved the undertaking of the Rhineland Study and it was carried out according to the recommendations of the “International Council for Harmonisation Good Clinical Practice” standards.

## Consent to Participate

Written informed consent was acquired from all participants per the Declaration of Helsinki in both the NEO and Rhineland Study.

## Data Availability

To protect participant privacy and comply with legal regulations, the NEO study data is not publicly accessible. Qualified researchers can request access by contacting the NEO Executive Board at https://www.lumc.nl/org/neo-studie/contact/. Similarly, the Rhineland Study data used in this manuscript is restricted to public access due to data protection laws. Researchers seeking access to these datasets can submit requests to, providing evidence of their qualifications and adherence to the respective study’s data use policies. All authors had full access to the data from their respective studies and are responsible for the accuracy and integrity of the data and analysis.

## Supplementary Figures

**Figure S1:**
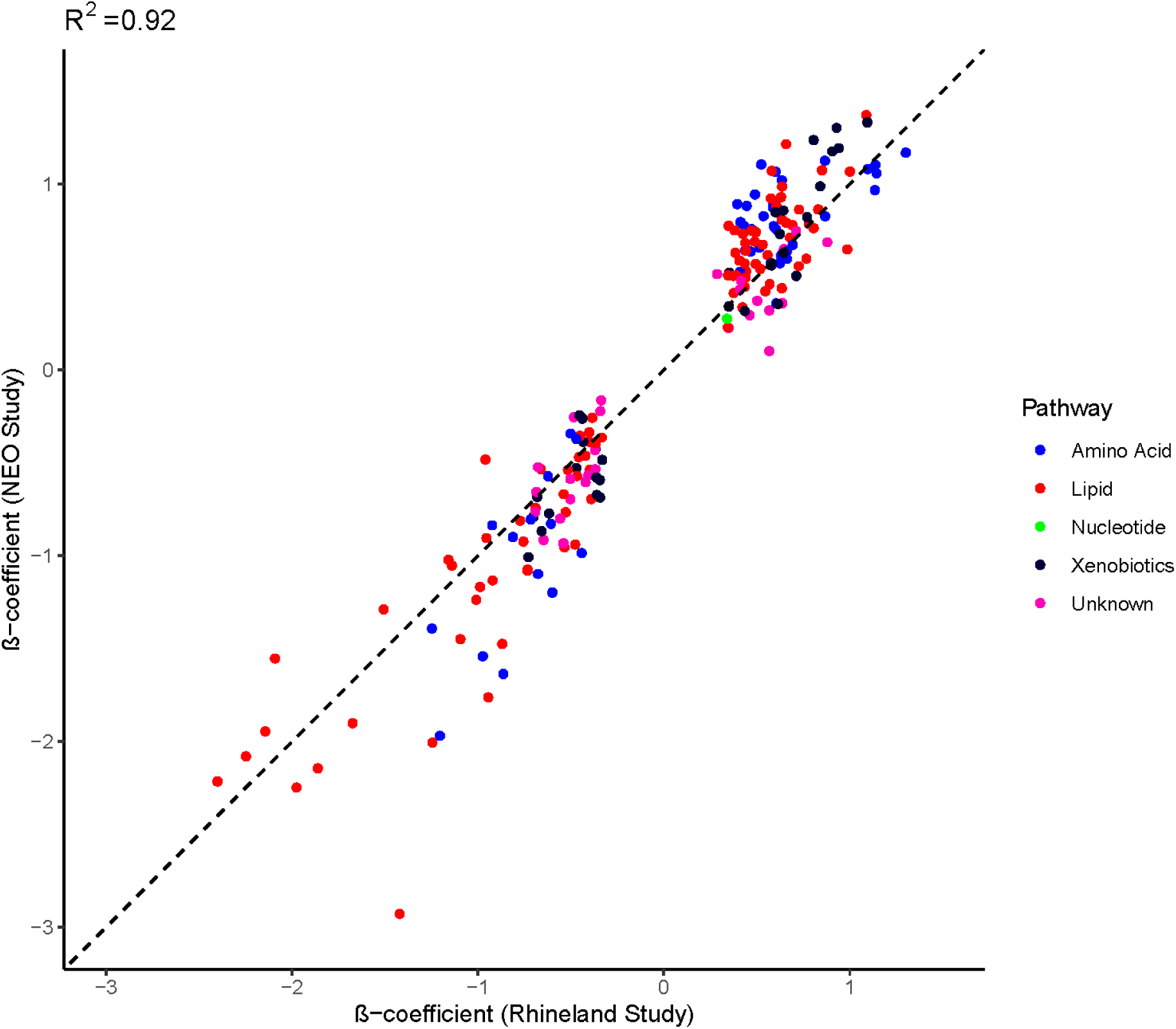
Correlation between the GWAS meta-analysis independent SNP(r^2^<0.6)-metabolite effect sizes from the Rhineland Study and the NEO Study.

**Figure S2:**
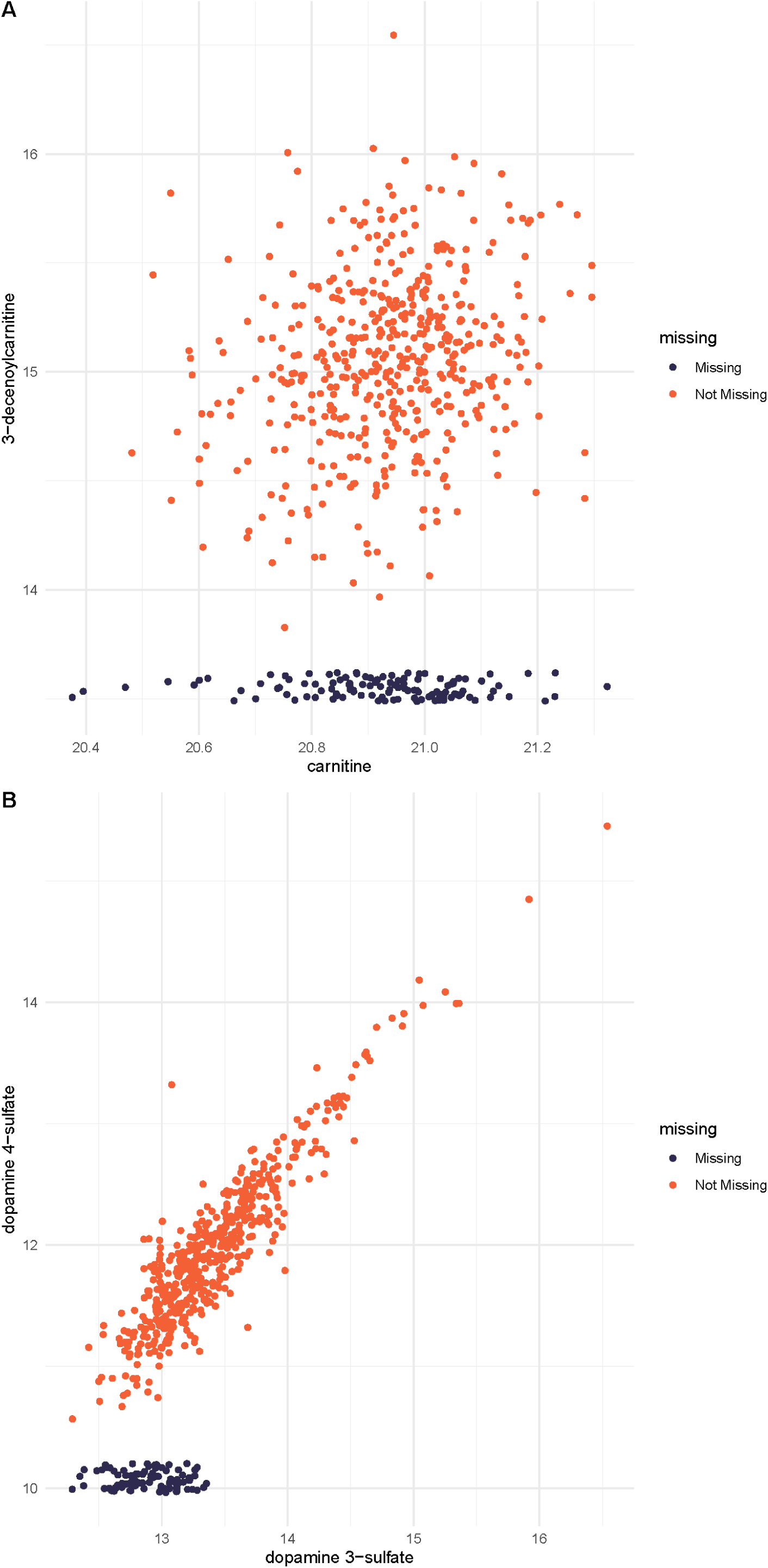
Correlation and visualization of missingness of 3-decenoylcarnitine and dopamine 4-sulphate in relation to carnitine and dopamine 3-sulphate in the NEO study.

**Figure S3:**
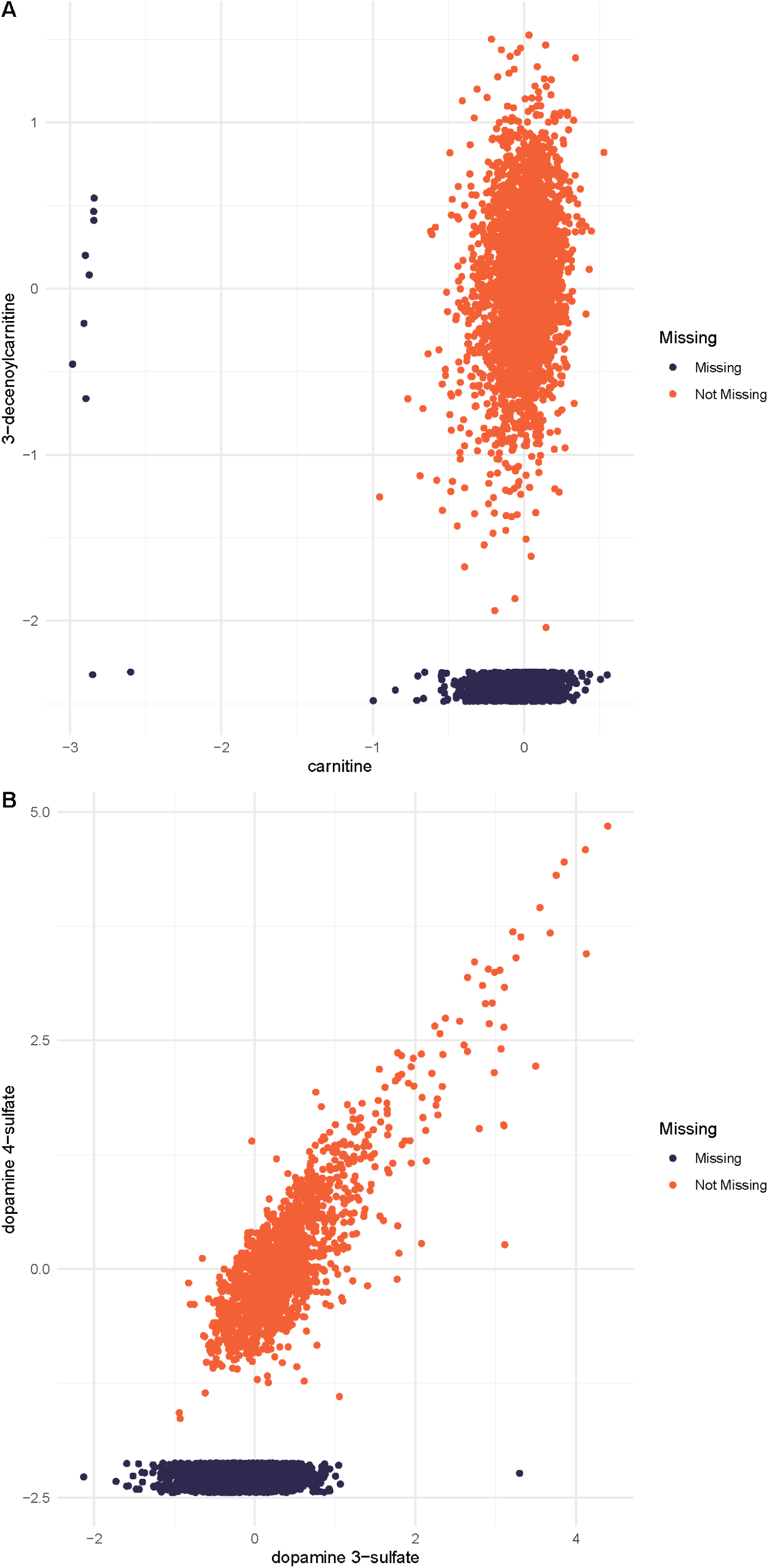
Correlation and visualization of missingness of 3-decenoylcarnitine and dopamine 4-sulphate in relation to carnitine and dopamine 3-sulphate in the Rhineland Study.

